# Strain variation and anomalous climate synergistically influence cholera pandemics

**DOI:** 10.1101/2021.04.07.21255051

**Authors:** X. Rodó, M.J. Bouma, M.A. Rodriguez-Arias, M. Roy, P. De Yebra, M. Garcia, M. Pascual

## Abstract

Explanations for the genesis and propagation of recurrent cholera pandemics since 1817 have remained elusive. Evolutionary change of the pathogen is presumed to have been a dominant factor behind the 7^th “^*El Tor”* pandemic, but little is known to support this hypothesis for preceding pandemics. We investigate the concomitant roles of climate and putative strain variation for the 6^th^ cholera pandemic (1899-1923; the one with the highest ever associated mortality in India), using newly assembled historical records for climate variables and cholera deaths for Bengal, Assam, Bihar and Bombay provinces in former British India. We compare this historical pandemic with the 7^th^ (*El Tor*) one and with the temporary emergence of the *O139* strain in Bangladesh and globally. Finally, we ran multi-model climate simulations to infer past and future long-term means of rainfall distributions on the basis of 39 models for 1861-2100, and for different periods of 50 years (1875-1925; 1975-2025 and 2050-2100).

The 6^th^ cholera pandemic featured a large scale synchronisation with a delay of a few years in both seasonal and interannual cholera variability over the endemic Bengal region during the El Niño event of 1904-07. Additional evidence supporting the establishment of a new strain includes a shift of cholera incidence to older age groups, an increase in the case fatality rate and the suppression of the spring cholera peak.

The 6^th^ cholera pandemic of Indian origin was associated with a novel and particularly invasive strain of new territory, and also with some delay, of endemic parts of India that act as a genetic regional reservoir of the disease. Climate anomalies appear to have played an important role in facilitating the establishment of this invasive strain, with environmental conditions similar to those underlying strain changes associated with ENSO in today’s Bangladesh. The evolutionary change of pathogens can act synergistically with climatic conditions in the replacement and propagation of emerging strains, as was the case in cholera’s 7^th^ pandemic. Increased climate variability and extremes under global warming would thus provide windows of opportunity for emerging new pathogens.

## 1. Introduction

The global spread of potentially fatal infectious diseases, such as plague, flu and cholera, had a significant impact on human history through large demographic and economic consequences. Although the clinical description of cholera is recorded in classical publications, and the disease has long been endemic in the Indian subcontinent, pandemic excursions of cholera from its endemic regions were only first reported in the 1820s. In the 19^th^ century cholera caused social shock waves reminiscent of those of plague in earlier centuries, striking regardless of age, sex and social class. Some strains of toxigenic *Vibrio cholera* resulted in explosive outbreaks when introduced into immunologically naive populations with poor sanitary infrastructure, as was evident in the devastating 2010 cholera epidemic in Haiti following the earthquake disaster^1-3^. Since the introduction of major and widespread hygienic initiatives for waste disposal and water supply during the 20^th^century, cholera has become a disease of poverty^4^, with an estimated 2.9 million of cases and 95,000 deaths in 69 endemic countries each year^5^

The origin of cholera and in particular the role of the environment has been debated for the last two centuries, long after the discovery by Koch in 1883 of its causative agent, the bacillus *Vibrio cholerae*^6^. On the one hand, the worldwide expansion of both the 6^th^and the 7^th^ pandemics has occurred in the form of several waves^7^, with the successive colonization of the different continents, allegedly following favorable climate and socioeconomic conditions in crowded low-income countries in Asia, Africa and ultimately, the Americas^8^. Thus, climate conditions would have facilitated the regional spread of the disease once seeded into a new area by human mobility. Human mobility in the context of climate variability has also been documented recently within the *El Tor* biotype for 1991 isolates from several countries (e.g. China^9^; Thailand^10^; and Angola^11^).

On the other hand, genetic changes of the pathogen associated with severe outbreaks and clinical and epidemiological manifestations have implicated evolutionary change in *V. cholerae* as responsible for previous pandemics^12-14^. Travel can fuel propagation of the bacterium under either scenario. Evidence of travel from cholera-affected cities as a source of the pathogen was presented for the large Danish 1853 cholera outbreak which killed 3.4%–8.9% of the population, with the highest mortality among seniors (16%) and the lowest among children (2.7%)^15^.

The 7th pandemic is specifically associated with the propagation and partial replacement in endemic parts of India and Bangladesh of the *Classical* biotype by the *El Tor* strain in the 1960s. Multiple introductions in Africa from its putative origins in the Bay of Bengal have also re-emphasised in recent years the human mobility component of pandemic cholera. The more recent appearance of the *O139* strain temporarily raised the prospect of a new cholera pandemic^16^. The isolation and identification of cholera only possible since the 1980s led to the classification of the biotype preceding *El Tor* as *Classical*, which would have been responsible for the 5^th^ and 6^th^ pandemics and most likely for the preceding ones^17^. The extensive genetic variation within the *Classical* biotype is likely to have fostered the 6^th^ pandemic. The exact origins and impact of the 1899-1923 pandemic in India remained however rather obscure due to over 800,000 cholera deaths in 1900 for India, and the Viceroy’s moratorium on related publications for fear of generating adverse publicity for the colonial enterprise.

To date, the interaction of climate conditions and strain variation has not been investigated as a driver of pandemic cholera. Increasingly available genomic and environmental data enable today consideration of these factors in ways never possible before. There is also growing interest in the potential contributions of both climate conditions and evolutionary change to the emergence of other pathogens, including for example chikungunya, dengue and more recently Zika^18,19^. In this study, we analyse extensive data from historical records on cholera mortality covering the onset of the 6^th^ and 7^th^ cholera pandemics, and present evidence for the relevance of both strain novelty and anomalous weather conditions. We end with potential implications for (pandemic) disease emergence and variability in a warmer world.

## 2. Data and methods

The monthly cholera mortality figures (1893-1939) and percentages of villages with reported cholera deaths were collated from British libraries based on the annual reports of the sanitary commissioners for districts in Bengal^20^, Punjab^21^ and Calcutta^22^. In addition to records for the non-endemic Bombay Presidency, these reports provide spatially explicit data for the other endemic Provinces traversed by the Ganges and Brahmaputra traversed by the 24 Bengal districts described in previous studies^23^ (see also the SI). Districts in Orissa and Bihar which were part of the Bengal Presidency before 1912, were excluded from the analysis. In addition, incomplete reports are available for case fatality rates of “boats” men living in the harbour (e.g. “the native floating population”) in Calcutta (Fig. 1, 1900-1912), and for cholera mortality in the Punjab by age group from 1908 to 1919 (Fig. S1A)

**Figure 1.**
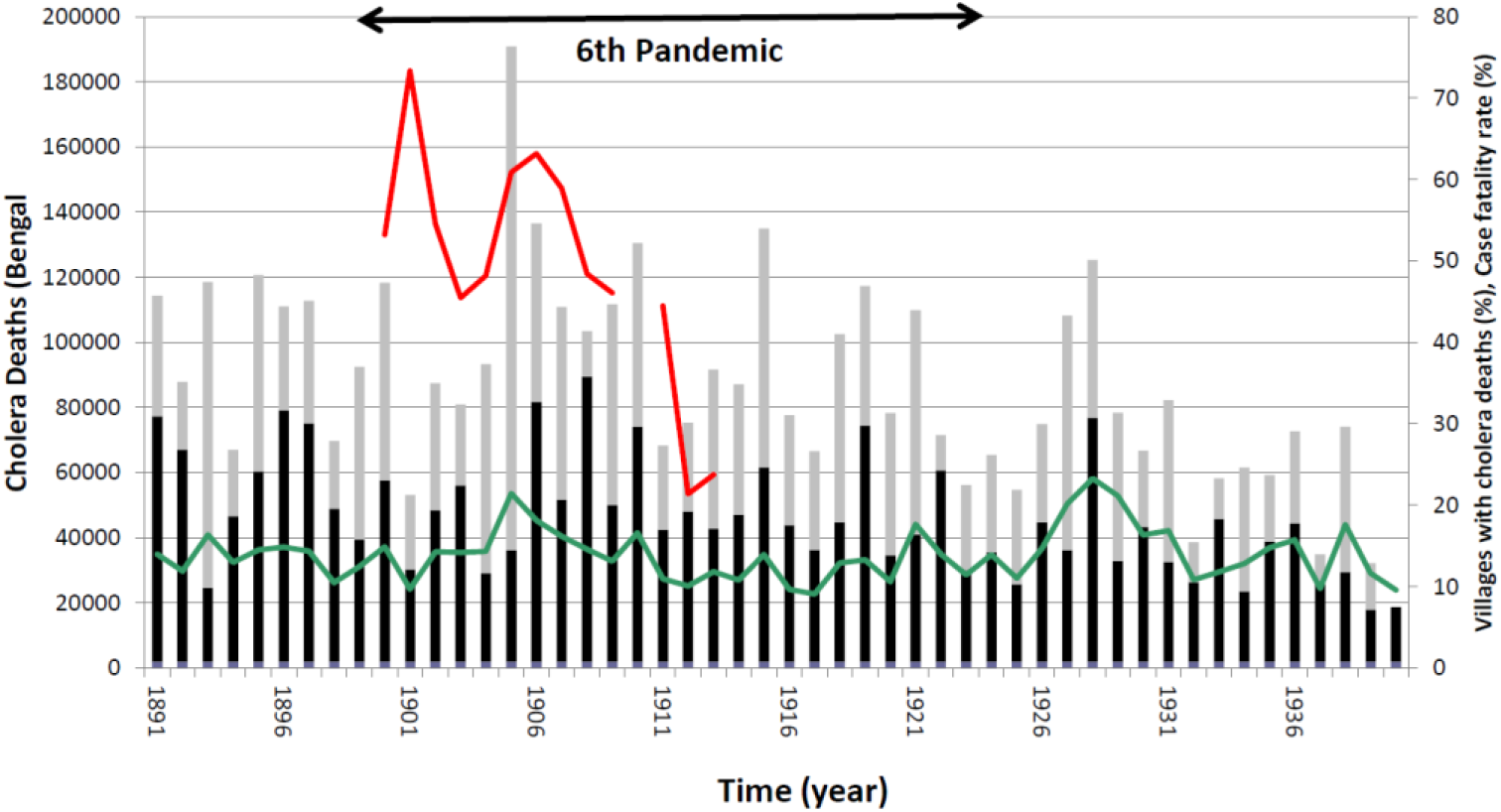
Cholera deaths in Bengal Province in the 6th pandemic. Deaths between February-July (black bars) and between August-January (grey bars). Case fatality rate (%) in Calcutta harbour, the “floating population” (Red) and the % of villages in Bengal with cholera deaths (green) on Secondary Y axis). Mortality (Bars). 24 districts of Bengal, with 86,353 villages (in % of villages with cholera deaths).

For the period of 1893 to 1939, climate data were obtained from different sources due to the lack of available products covering both the 19^th^ and the 20^th^ centuries together. Rainfall and minimum and maximum temperature from surface station data were retrieved from the historical archives of former British India^24^ (see also the SI). For the historical period, we relied on sea-surface temperatures (SST) anomalies from the Twentieth Century Reanalysis (TCR). We also generated averages from land station values for both monthly temperature in Dacca (°C) and the North East India Rainfall (NEIR, mm) from the climatic database of the Indian Institute for Tropical Meteorology at Pune (http://www.tropmet.res.in). In addition, to represent rainfall in more recent Bangladesh spatially, we used the gridded rainfall data from Global Precipitation Climatology Project (GPCC)^25^. For rainfall time series, NEIR regional precipitation was preferred to Dacca’s local rainfall as it represents a better way to characterize the total water accumulation in the catchment area of interest.

The Niño3.4 SST index of ENSO over coastal south America was considered to address associations of cholera with this major driver of global climate variability (http://www.esrl.noaa.gov/psd/people/cathy.smith/best/).

The synchrony of cholera mortality in the different districts was evaluated with wavelet coherence spectra, computed from the wavelet power spectra for the two individual time series (WPS)^26,27^. This approach was selected to specifically examine associations that are transient in time and to analyse timeseries that are themselves non-stationary. Under such conditions, the classical Fourier power spectrum can be problematic since it averages over time the contributions of the different frequencies, and therefore these contributions cannot be localized in time. In contrast, the wavelet spectrum provides a decomposition of the variance of a time series into different frequencies explicitly over time. Similarly, the resulting coherence spectrum indicates not just whether, but when in time, two time series exhibit variance at similar frequencies. (For detailed description of these methods and application to epidemiology^28^).

Scale-Dependent Correlation Analysis (SDC^29,30^) was applied to examine correlations that are local in time between the different cholera strains and ENSO. Also, Multichannel Singular Spectrum Analysis (MCSSA^31-32^) was applied to decompose cholera variability into major (orthogonal) components accounting for the largest amount of its variance, and for time series reconstruction based on given components (see 33 for an explanation with an application to epidemiology). Here, reconstructions are specifically obtained by removing the trend and keeping all other components, including seasonality and interannual variability.

Climate variability in both space and time was decomposed via Principal Components Analysis (PCA) to identify, and discriminate among, main modes of variability in a particular variable, and then address associations of the dominant temporal modes (Principal Components, or PCs) with other variables (cholera levels^34^). The resulting empirical orthogonal functions (EOF) provide dominant spatial components over which temporal variation in the form of PCs can be examined.

Finally, to assess the degree of severity of the climate anomalies that accompanied the historical epidemiological events, and to establish expected changes in the future under climate change, multi-model climate simulations were performed for the relevant region (87.5 to 92.5 deg. longitude and 20 to 27.5 deg. latitude corresponding to current Bangladesh). Specifically, one-thousand multi-model simulations based on the Coupled Model Intercomparison Project Phase 5 (CMIP5) were averaged over the region for three 50-yr time periods (1875-1925; 1975-2025 and 2050-2100). The future scenario considered here is the one known as RCP8.5 which represents the worst-case scenario for global greenhouse gases emissions and the one the world is currently experiencing. (For further details see SI).

(For readers not familiar with these statistical methods we provide additional description in the SI and we guide interpretation of particular results in the text below and figure captions).

## 3. Results

The 6^th^ cholera pandemic commenced with a dramatic surge of cholera deaths in British India, its endemic “homeland”. In 1900 an unprecedented 700,000 deaths were reported from the provinces. In the Bombay Presidency, the low baseline rate from around 1 cholera death per 1000 increased nine-fold. Mortality was not above normal in Bengal, the most endemic province in 1900 (Figure 1), but a very steep rise in the case fatality was apparent from the harbour population in the capital Calcutta, suggestive of the introduction of a new strain (Figure 1). Only in 1905 cholera deaths in Calcutta rose sharply to over 180,000, two standard deviation above the mean, and the fraction of villages in Bengal with cholera mortality rose over the whole vast deltaic region (Figure 1)^35^. Also the case fatality of the “floating population” rose again in 1905, before settling in 1912 to the normal rate of about 20% (Figure 1). Strikingly, the vast seasonal variation in cholera mortality between the 24 districts of Bengal vanished in 1905, with a marked development of synchrony and a shift of deaths to the fall (Figures 1 and S1B,C).

The synchrony of this change in mortality between districts in Bengal is shown in figure 2A. The wavelet coherence spectrum was computed between monthly cholera deaths in Dacca (used as reference) (Fig. 2A-B) and individual districts along the Brahmaputra basin (i.e. pink districts in Fig. 2B). Typically, the two coastal districts display a distinct seasonality^36^. But around 1905 all district pairs exhibited full synchronization. This spatio-temporal coherency is evident for the annual cycle (12 months in the vertical scale, Fig. 2A), indicating a temporary amplification of the seasonal cholera cycle.

**Figure 2.**
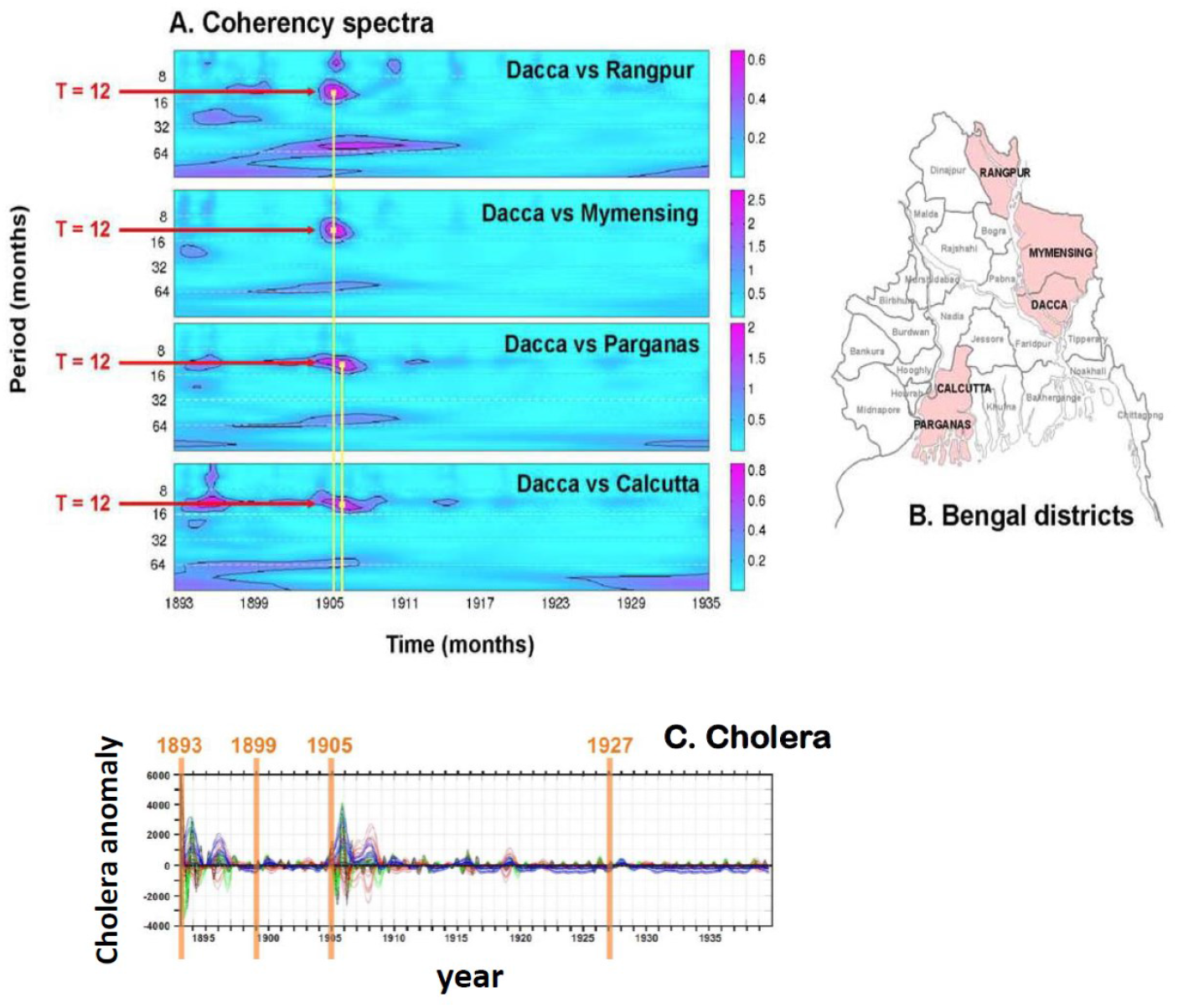
Cholera synchronisation in Bengal. A) Wavelet coherence spectra between monthly cholera mortality in Dacca from 1893 to 1935 and that of selected districts in the different regions of former Bengal (Rangpur, Mymensing, Calcutta and Parganas). Each plot shows the intensity of the coherence as a function of time (in the x axis) and period (in the y axis). Areas in the plots with high power indicate synchronization. (Areas were the signal is significant are surrounded by black lines). Red horizontal lines denote the time scale (the period), and the yellow vertical lines, the time when synchronization took place (1904-07). B) Map of the Bengal districts at the beginning of the 20^th^ century with coloured districts indicating those used for the cholera analysis. C) Seasonal-to-interannual components of cholera for all districts in Dacca for the interval 1893-1940, reconstructed with Singular spectrum analysis (MCSSA) (*see SI methods for description*). Note the two times (1893 and 1905) around which the variability of cholera anomalies at all temporal scales is more pronounced, denoting an intense alteration of the normal pattern.

Reconstructed components covering all time scales for cholera except trend generated with Singular Spectrum Analysis (SSA^31, 32^) for each time series in Bengal, lend further support to the anomalous synchronisation across the region (Fig. 2C). The less striking synchronisation in 1893 (Figure 2A), and the increased mortality in 1927 in the absence of synchronization (Figures 1, 2C) may reflect strain diversification with less dramatic consequences.

We interpret this spatial synchronisation in this endemic region as a manifestation of a new strain of cholera for which the population lacked protective immunity. These changes in 1905 resemble indeed those in the 1960s, when the *El Tor* strain replaced the “*Classical*” on its march to global hegemony; with similarities extending to a delay in strain replacement, a shift of seasonal deaths to the fall period (as described below) and a raised case fatality rate. Data to investigate a shift of morbidity and mortality to an older age-group, signifying failure of acquired immunity, documented during *El Tor* and *O139* strain replacements were not available for Bengal. However available data by age and sex (1908-1919) for the Punjab province in NW part of the subcontinent show such a shift in 1911 (Fig S1A). Apart from the ratio between cholera mortality in the population over and under 10 years of age increasing five-fold, an increase in the male-to-female ratio (>10 years) has been observed in non-endemic areas struck by a cholera^, 38^, and is presumably related to a higher exposure of the male population.

The noticeable delay between the apparent introduction of a new virulent strain and its ability to outcompete the incumbent one, suggests other determinants may have been at play. Weather is a prime potential candidate we investigate next. Fig. 3A summarizes the anomalous seasonal patterns of mortality, rainfall and temperature in Dacca during the exceptional period of 1904-07. In contrast to the typical bimodal seasonal pattern of the disease with two peaks per year, the anomalous seasonality was characterized by a single, but extremely large, annual cholera peak (Fig. S1C; Fig. 3A). Specifically, the spring peak was absent and the winter cholera peak grew in conjunction with decreasing rainfall after heavy and delayed monsoon rains in an environment where temperature is suitable for cholera proliferation. These conditions typically lasted until December when temperatures fell below approximately 21 degrees. Anomalous high cholera incidence occurred during the three winters, particularly in 1904/05 and 1905/06 (Fig. S1C), following anomalous monsoon rains, initially lower than normal for June (1904 to 1906), and extremely high values in August 1905 (*with more than 150mm excess rain*) and August 1906 (*over 75 mm excess rain*). These monsoon seasons were also preceded by anomalous spring conditions in both rainfall and temperatures. February 1905 experienced extreme low temperatures (Fig. S1C and Fig. 3C), more than 3 degrees below the seasonal average, a situation that did not return to normal until late spring. In conjunction with this severe decrease in temperatures, a concomitant abnormal increase in rainfall occurred in March 1905 (Fig. 3B2) with absolute values more than double the typical average.

**Figure 3.**
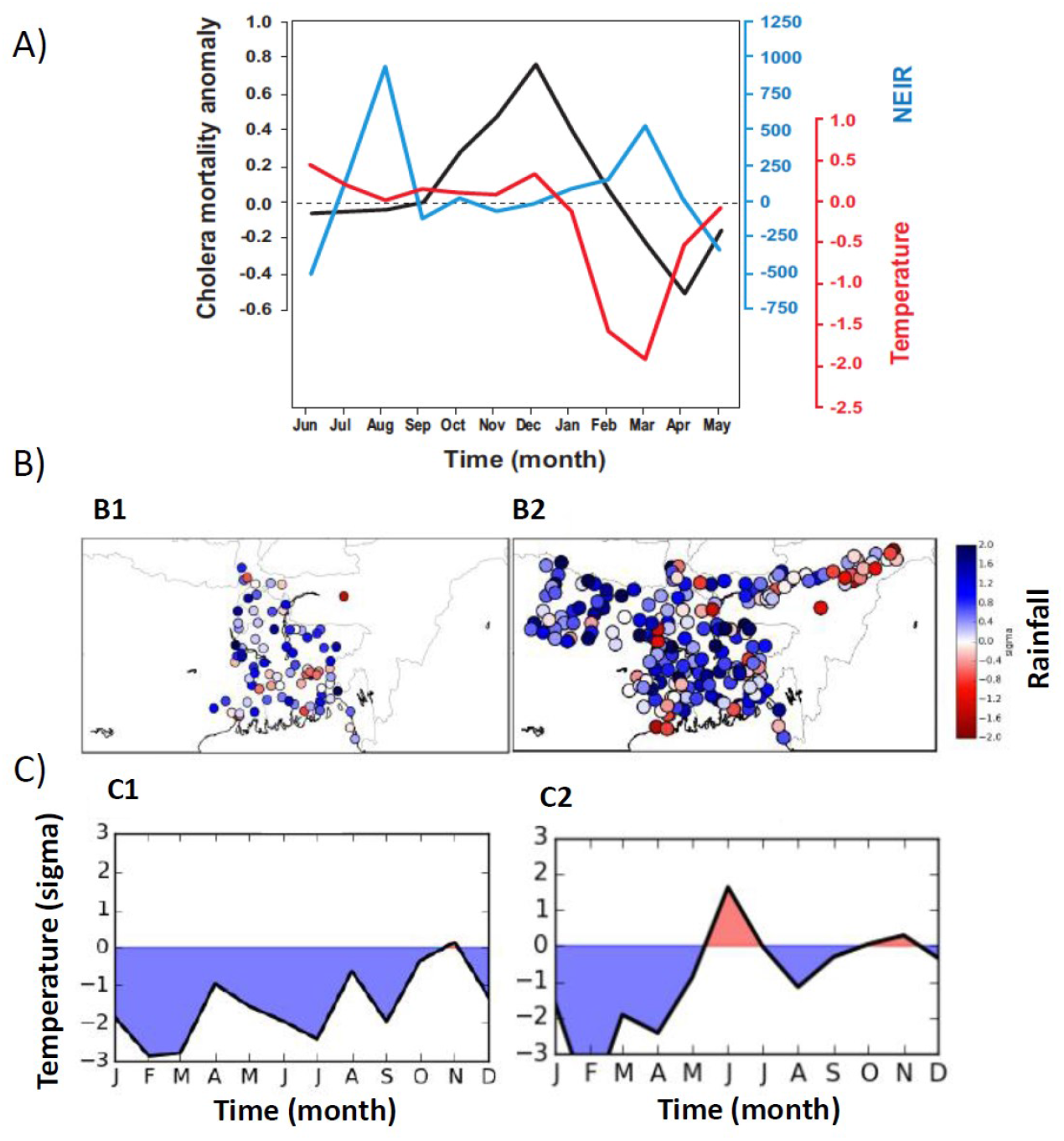
Cholera, rainfall and temperature anomalies in the 1904-05 event. A) Difference between the average of the 1904-07 event and the seasonal cycles of cholera mortality anomaly (black), rainfall from NEIR (blue) and temperature (red) in Dacca. B) Rainfall anomalies for Bengal in 1893 (B1) and Bengal, Bihar and Assam in 1905 (B2). Colors denote the number of standard deviations from the mean of rainfall in the intervals 1893-1939 (B1); and 1901-2010 (B2). Precipitation anomalies are shown for JJAS. B3 and B4 display temperature anomalies (deg. °C) as standard deviations from means in the respective interval years.

Comparison of the 1904-07 event with normal years for climate conditions (Fig. 3A and Fig. S1B,C) highlight the delayed and more intense monsoon, with the rainfall maximum in August, and a long winter characterized by low temperatures that last until April. Among these months, anomalous higher than normal rainfall is again observed in February and March 1905. Exactly the same conditions are experienced over most of the country in 1905 (Fig. 3B2, 3C2), with very cold anomalies already beginning the previous winter and with an intense and delayed monsoon. These same patterns occurred during the extreme cholera anomaly of 1893 (Fig. 3B1, 3C1).

Rainfall variability over the region for 1901-1939 exhibits a marked spatial structure as revealed by Principal Component Analysis (PCA) of the GPCC gridded reanalysis data (Fig. S2A). The two main spatial components (the first and second Empirical Orthogonal Functions, EOFs) for rainfall display a NE-SW axis of variability identifying a dominant pattern either along the Ganges (NW-SE) or the Brahmaputra (NE-SW; Fig. S2) respectively. The first temporal PCs (PC1) corresponding to the main spatial mode (Fig. S2B), indicates a high rainfall score over the whole country for 1905. Highly anomalous cholera years are indicated as red dots. Moreover, both PC1 and PC2 are significantly correlated with ENSO (*p<0*.*01*), consistent with a strong relationship between rainfall and ENSO known in the literature to be stable at least until late 1970s^39^ (when an ocean regime shift occurred in the Pacific). Therefore, high scores in PC1 might also be related to large ENSO variability according to these analyses.

The years of 1893 and 1905 correspond to the two major ENSO anomalies (in absolute terms) occurring in the interval of 1880-1940, namely a La Nina event in 1893 and an El Nino one in 1905 (Fig. 4A). ENSO-related teleconnections in rainfall and temperatures have been associated in different parts of the world to more recent temporal patterns of cholera^40-43^. Specifically, cholera cases during the global expansion of the 7^th^ pandemic can be related to the facilitation by ENSO’s regional associated teleconnections in Asia (1963-64), Africa (1991-92; 1997 and 2002) and the Americas (1991-92). Fig. 4B provides a summary of these temporal and spatial links with ENSO events and places the 1893 and 1905 El Niños in context^44^.

**Figure 4.**
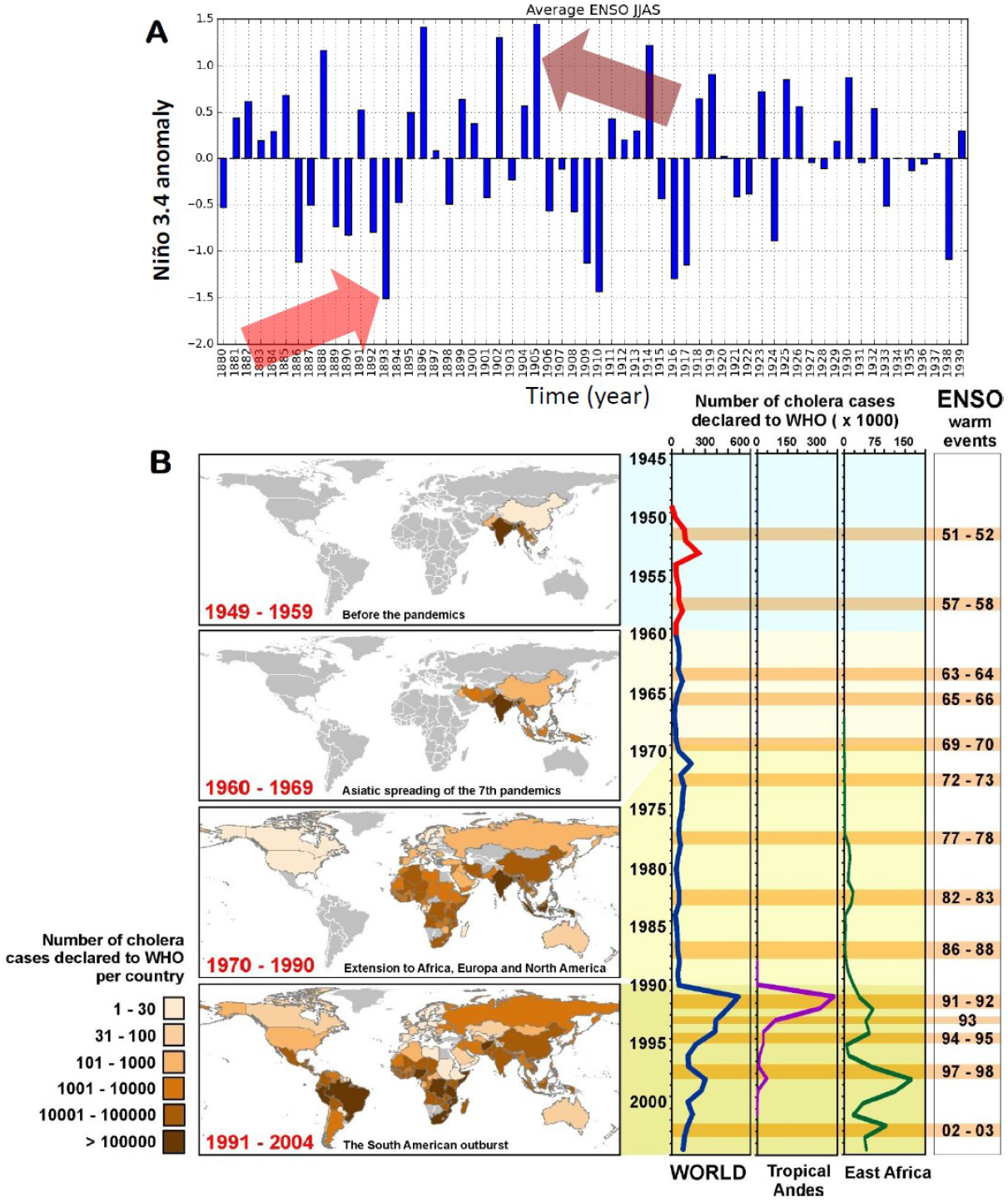
El Niño-Southern Oscillation and the global expansion of cholera. A) ENSO index data from 1880 to 1939 retrieved from http://www.esrl.noaa.gov/psd/people/cathy.smith/best/. Arrows indicate locations of the 1893 and 1905 strong ENSO events. B) Left: Global propagation of cholera in the 7th pandemic (1950-2004). A) Geographic spread showing colonisation of different continents in terms of cholera cases reported by country to WHO. Right: Number of accumulated cholera cases (x1000) in the world, tropical Andes and East Africa, and the successive El Niño events on record.

For comparison, we also examined the contribution of ENSO to cholera’s exacerbation at times of strain replacement in present-day Bangladesh. To investigate ENSO’s influence on regional climate, Scale-Dependent Correlation Analysis (SDC)^45^was applied between global sea-surface temperature (SST) anomalies and ENSO (i.e. Niño 3.4 index). The resulting maps show moving-window correlations calculated between the Niño 3.4 index and the SST series at each grid point of a 2.5 degrees’ resolution of a global database. These maps illustrate the anomalous oceanic configurations occurring during the three El Niños (EN), including a strong (1987) and very strong (1982 and 1997) events known to have operated distinctly over the Indian subcontinent^46,33,39^ (Fig. S3A). Specifically, global ocean SSTs exhibited changes during these EN events consistent with impacts over the Indian subcontinent.

These changes occur at times that are associated with cholera events as shown in Fig. S3B (*right panels*). This can be seen in the SDC analyses^3340^ now applied to correlations between ENSO (Niño3.4 index) and total cholera cases, individually for each of the three different strains in Bangladesh (Fig. S3B for *Classical, El Tor* and *O139, a variant of the El Tor strain*). In all cases, ENSO led cholera by between 7 to 12 months, a delay consistent with previous studies^40,47^. Strikingly, local monthly correlation maxima cluster around the years when cholera experienced the strains replacements, as well as the large increases in cases for each of the three strains. In particular, these clusters are clearly found for the *Classical* strain following the very strong El Niño event of 1982, for *El Tor* replacing *Classical* in 1986-88, and for *O139* overtaking *El Tor* from1994 to 1997.

We also analysed the onset of the 7^th^ pandemic and its relation to climate anomalies (Fig. S4). Despite the *El Tor* biotype (serogroup O1) being first identified in 1905 in a quarantine camp in *El Tor* (Egypt) which gave it its name, and then identified again in 1937, the pandemic did not arise until 1961-62 in Sulawesi and Semarang in Indonesia. The expansion to the entire Indonesia took place in 1962 and then to Bangladesh in 1963 and India in 1964. As seen in Fig. S4A, 1961, the year of cholera’s appearance in the two above locations in Indonesia was not anomalous in terms of the absolute rainfall, although 1961 was a drought year over the archipelago (*not shown*). Instead, 1962, the year of cholera’s spread in Indonesia and beyond was characterised again by very cold temperatures and above-rainfall conditions region wide (Fig. S4B), with anomalies of over 2 standard deviations registered in both variables. In other words, the climate anomalies occurred with some delay, following the timing of the first emergence.

Finally, to assess the degree of severity of the 1905 positive rainfall anomalies during the delayed monsoons, and to consider the expected changes in the future under the current evolution of greenhouse gases emissions (known as +RCP8.5 scenario), the results of multi-model climate simulations for the region are shown in Fig. 5. Comparison to the distributions for three 50-year periods (past:1875-1925; present:1975-2025 and future:2050-2100) shows that the conditions experienced during 1904 and 1905 were very extreme (arrow), lying at the tail of the distributions for past and future simulations. Also, distributions of rainfall shift towards more intense values for the future climate change scenario.

**Figure 5.**
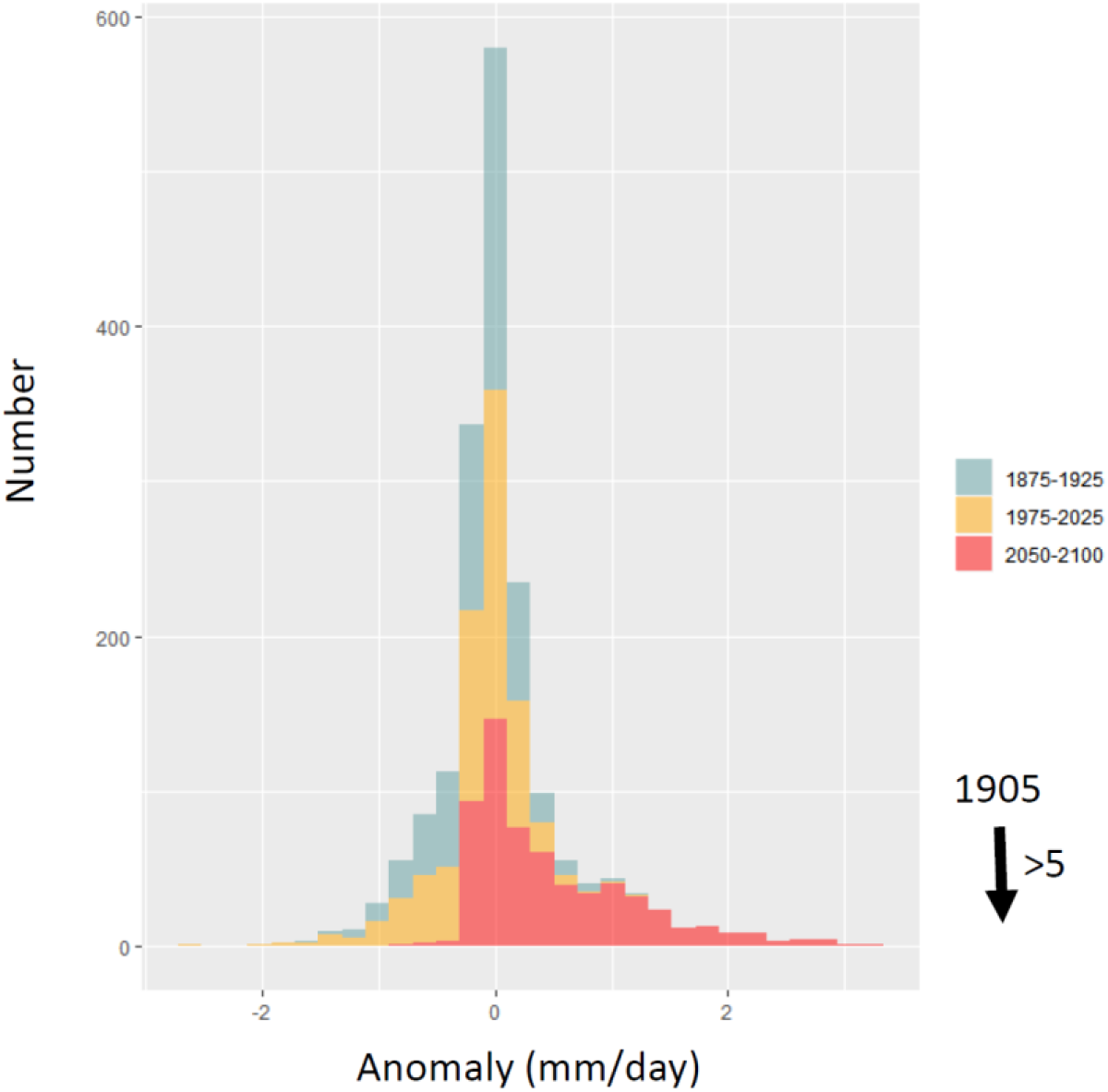
CMIP5 multi-model climate simulations for rainfall in Bangladesh. Distribution of rainfall anomalies in [mm/day] from CMIP5 multi-model simulations averaged over the region (87.5 to 92.5 deg. longitude and 20 to 27.5 deg. Latitude), corresponding to current Bangladesh. The distribution denotes the number of times a simulated value is obtained in 1000 model runs. For each model output, the points considered were only those where the corresponding station data had more than 50% of rainfall measurements present. The data shown correspond to the multi-model mean of historical (+rcp85) experiments. Arrow indicates for comparison the value of the 1905 rainfall anomaly. The list of climate and earth system models is given in the SI.

## 4. Discussion and conclusions

The pandemic excursions of cholera remain among the least understood phenomena in infectious diseases research. The unique complete synchronization event described here for Bengal between1904-1907 suggests a major external force was at play on the otherwise spatially differentiated cholera dynamics of this region. One possible explanation for such a synchronous and extreme cholera event is climate anomalies acting over a large geographical region (i.e. Moran effect^48,49^). Another explanation is the emergence of a novel strain. Acting on top of a secular trend in mortality (Figure 1) explained by improvements in treatment and sanitary conditions^23^., the introduction and full establishment of a new and possibly more virulent strain would result in higher mortality at the regional scale. As documented for the most recent succession of strains^37,38^, an increased mortality due to strain emergence would be accompanied by a relative shift in mortality to older age groups as documented for the most recent succession of events^37,38^. This shift reflects a lack of immunity in age groups with protection against the previously dominating strain.

The hypothesis that strain variation has been responsible for the recurrent changes in apparent virulence associated with pandemics^50^ is difficult to test, as strain identification only commenced after isolation of the bacterium during the 5^th^ pandemic. By using other epidemiological characteristics of the appearance of new strains in recent decades, we presented evidence consistent with a novel strain in the initial stage of the 6^th^ pandemic. A gap exists, however, between the first recognised outbreaks of the 6^th^ pandemic around 1900 (Calcutta and Bombay) and the highest and synchronous all India cholera mortality in 1905. This delay does not necessarily contradict that these two events, first appearance and emergence, are related. A new strain might have made its first appearance in 1899-1900, becoming fully established and replacing the previous strain a few years later. Such a sequence of events was indeed observed for the replacement of the *Classical* strain by *El Tor* in the same area at the beginning of the 7^th^ pandemic. *El Tor* accounted for a small proportion of cases in a Dacca Hospital between 1968 and 1972, and it completely replaced *Classical* in 1973 for a number of years^51^. After some time, there was a reoccurrence of the *Classical* strain and the apparent spatial differentiation of both strains in Bangladesh^52^. Similarly, *El Tor’s* genetic shifts occurred in 1992 in China related to the replacement of the prototype strain from 2002 to 2010^9^. Altered variants of atypical *El Tor* biotypes were also identified in India and Mozambique and completely replaced former cholera isolates in Thailand^10^, Vietnam^53^ and Angola^11^ around 1991. For all those years and places, El Niño impacts were documented in previous studies, consistent with the notion that altered climate variability favours cholera epidemics. Recent large-scale disease events took place regionally during periods of time that encompassed strong El Nino (EN) years, as noted for South America during 1991-92^8^ and for the large outbreaks in Africa after 1997/98 and 2002/03. Disease synchronization among regions affected by strong EN events has also been documented for the latter outbreaks^41^.

For the historical data from former British India, we relied here on these known features of changing strains to retrospectively assess the evidence for a strain shift during the 6^th^ pandemic (1899-1927). The unique spatial resolution of these records allowed us to examine cholera’s synchronization over an unprecedented large region, across districts, and in the context of a long temporal record.

There was no shift in the seasonality of cholera after 1905, a pattern observed later when *El Tor* replaced the *Classical* strain^51^ with a well-documented winter peak shift from December to mid-October, and also when *O139* appeared in the 1990s. Differences in seasonality between strains, most likely reflect different ecological requirements between strains^50,54^ that do not necessarily accompany antigenic and virulence changes. We do document a higher adult male mortality in the Punjab a few years after the 1904-07 event, a characteristic of the more recent replacements. Similar changes in epidemiology were reported in former strain shifts (i.e. the Danish cholera outbreak of the 1850s^15^). These include a change in virulence also reflected in the case-fatality rate (for *O139*) and a shift of the disease burden to an older population (*El Tor* and *O139*), associated with the lack of specific immunity to the new strain.

Several of our results support a role of climate acting as a major driver of the 1904-07 anomalous cholera episode, which would have facilitated the establishment of the novel strain. The atypical suppression of the cholera spring peak during the 1904-07 abnormal cholera interval was shown to be related to anomalous cold conditions and high precipitation. In addition, higher mortalities in the dominant winter peaks would have been facilitated by the anomalous monsoon season of the previous summer. Thus, different drivers appear to underlie the two cholera peaks in a normal year, consistent with studies of environmental factors determining the seasonality of the disease. Very low temperatures in late winter would have helped in sharply ending the winter cholera peak. Similarly, low temperatures continuing through mid-spring (of the order of two to three degrees lower than normal) and concurrent high relative rainfall (an excess of more than 1,000 mm) might have led to the full suppression of the spring cholera peak that normally follows. Temperature during these months has been shown to affect the intensity of the spring peak^23^. Both effects would have generated a larger pool of susceptible individuals or/and lower levels of temporary acquired immunity, contributing to a large winter outbreak in the transmission season that followed the monsoons. Extremely heavy rains in the monsoon season may also have played an important role by disrupting sanitation systems and promoting with a delay the proliferation of bacteria in the environment due for example to increased nutrients. Evidence for a dual role of rainfall in the seasonal cycle of cholera with both a negative and a positive effect at different lags has been presented for historical cholera in endemic regions of Madras^54^ and former Bengal^55^. Anomalous rainfall conditions can also affect harvest levels, enhancing population malnutrition and promoting famine and disease^56^.

The spread and extent of the 7^th^ pandemic is presumed to be due to the relative mildness of *El Tor* relative to *Classical* which leads to more asymptomatic individuals (50 to 1 symptomatic case), as well as to the enhanced persistence of the bacterium in the natural environment^47^. According to WHO records at that time, spread was promoted by a combination of a long dry spell in Kuching where wells contained little water, and a sports event with people flocking into the town. The lack of a piped water supply led to people gathering around wells contaminated with faecal water^6^. Dry spells much more extreme than the one in 1961 were recorded however many times prior and after that year and similar contagions did not occur (Fig. S4A). In contrast and consistent with our findings for the earlier pandemic, Indonesia experienced in 1962 an extremely cold spell (below 2 standard deviations) that was long lasting (see Fig. S4B), as well as very rainy (see January through March in Fig. S4C).

In relation to the described regional anomalies in rainfall and temperature, several of our results also support a role of ENSO during the 1904-07 event, consistent with studies of more recent cholera and climate variability^8,36,33,40,41^. The strong quasi-quadrennial component of the mortality (with a 4-yr periodicity) in the 1899-1905 interval observed for cholera (Fig. 2A) could reflect the strong coupling of ENSO to monsoon dynamics, as indicated by other studies^57-59^.

In summary, our results indicate that genetic shifts in the dominant *Classical* strain together with facilitation by climate underlie the establishment and expansion of the 6^th^ (and 7^th^) pandemics. These concomitant effects have implications for the potential future emergence of other pathogenic strains, as they underscore the role of environmental conditions in generating windows of opportunity for the colonisation by novel variants. Recent observed changes in cholera strains already include the shift towards *El Tor* dominance and the several failed attempts by *O139* to replace *El Tor*. Interestingly, strong El Niño conditions co-occurred with successful strain changes in the last decades of the 20th century^33^. For the earlier 6^th^ pandemic studied here, it is interesting that during the two EN years of 1899 and 1905 most inter-hemispheric and global indicators of global variability manifested an intense peak in activity^59^.

Finally, we specifically demonstrated the extreme nature of climate conditions around 1905 by placing these in the context of monsoon rainfall distributions generated by climate models. Those conditions proved extremely rare for the past and also for the future under a climate change scenario. This should not be interpreted as similar magnitudes of anomalies being needed for emergence. More likely climate conditions that enhance transmission during particular windows of time and several consecutive years can act to facilitate emergence in cholera but also in other pathogens that are water-borne and vector-borne and therefore closely connected to the environment. The increased capacity to monitor genetic changes of pathogens should be coupled to advances in climate studies of infectious diseases including climate modelling and prediction, to provide warnings about potential emergence. The finding of a possible delay between initial detection and actual emergence indicates the relevance of climate conditions can extend beyond the former. For cholera, the shift of the distribution of monsoon rains over Bangladesh towards higher values suggests more frequent conditions for transmission and emergence. The synergy of strain variation and anomalous environmental conditions arising more often, provides a warning of continued and enhanced opportunities for emergence of the disease given the strong climate change signals in its homeland and beyond.

## Data Availability

All climate data is freely available from public repositories. Cholera datasets are available under justified request to Menno Bouma.

http://www.tropmet.res.in

http://www.esrl.noaa.gov/psd/people/cathy.smith/best/

ftp://ftp.dwd.de/pub/data/gpcc/html/fulldata_v6_doi_download.html

https://www.rdocumentation.org/packages/Hmisc/versions/4.2-0/topics/rcorr

## 5. Acknowledgements

We thank the Indian Institute for Tropical Meteorology at Pune (http://www.tropmet.res.in) for supplying meteorological data and grants by NSF-NIH (Ecology of Infectious Diseases) and NOAA (Oceans and Health) to M.P. X.R acknowledges the support of the PERIS-PICAT project of the Catalan Dep. Salut and PARA-CLIM-CHANDIRGARGH of the New Indigo EU-India program. We acknowledge support from the Spanish Ministry of Science and Innovation through the “Centro de Excelencia Severo Ochoa 2019-2023” Program (CEX2018-000806-S), and support from the Generalitat de Catalunya through the CERCA Program.

## Author’s Contributions

X.R., M.J.B. and M.P. conceived the study. X.R. led the design and implementation of statistical analysis, M.J.B. recovered historical data for cholera and climate British India and contributed to data analysis. M.P. participated in the design and statistical analysis. M.G., P. de Y. and M.A.R. retrieved data and also performed analyses. M.P. and X.R. took the lead in writing the article. All authors discussed epidemiological interpretation and contributed to the writing.

## Supplementary Information

### Datasets

Historical cholera incidence datasets were retrieved from the annual reports of the Sanitary Commissioner for 1900 and 1901 in Bengal^20^ and Punjab^21^ and in Calcutta^22^)

In order to also cover the recent period, the gridded rainfall data from Global Precipitation Climatology Project (GPCC)^25^ were used for Bangladesh (retrieved from ftp://ftp.dwd.de/pub/data/gpcc/html/fulldata_v6_doi_download.html). These data are available at a 0.5° resolution for the whole 20^th^ century (1901-2012), and have undergone a strict homogenisation and quality control procedure to remove abnormal outliers and station biases. To assess whether the extended dataset in the Global Precipitation Climatology Project (GPCC^25^) can replace station rainfall for larger spatial domains than those of single stations, correlations were calculated between the first two PCs of the rainfall station data and those of the GPCC reanalysis database. Correlations are both significant (rPC1= 0.79 and rPC2= −0.67, respectively, *p<0*.*005*), indicating that the GPCC dataset is here a sufficiently good analog of station rainfall. The Twentieth Century Reanalysis (TCR) dataset for the region was also analysed for variables that can help better track climatic influence. Among these, sea-surface temperatures (SST) serve to better visualize weather dynamics related to local rainfall.

The Niño3.4 SST index of ENSO was considered to address associations with cholera (http://www.esrl.noaa.gov/psd/people/cathy.smith/best/). Niño 3.4 represents the sea surface temperature anomalies over a small region of the tropical Pacific near the coast of South America.

### Climate and earth system models used for the historical (+recp85) experiments

The following climate and earth system models were used in the multimodel RCP8.5 experiment: ACCESS1-0, ACCESS1-3, bcc-csm1-1, BNU-ESM, CanESM2, CCSM4, CESM1-BGC, CESM1-CAM5, CMCC-CM, CMCC-CMS, CNRM-CM5, CSIRO-Mk3-6-0, EC-EARTH, FGOALS-g2, FIO-ESM, GFDL-CM3, GFDL-ESM2G, GFDL-ESM2M, GISS-E2-H, GISS-E2-H, GISS-E2-H, GISS-E2-R, GISS-E2-R, GISS-E2-R, HadGEM2-AO, HadGEM2-CC, HadGEM2-ES, inmcm4, IPSL-CM5A-LR, IPSL-CM5A-MR, IPSL-CM5B-LR, MIROC5, MIROC-ESM, MIROC-ESM-CHEM, MPI-ESM-LR, MPI-ESM-MR, MRI-CGCM3, NorESM1-M, and NorESM1-ME. The interval of years for the long-term mean was computed from 1861-2100, and distributions are shown for three periods of 50 years (past:1875-1925; present:1975-2025 and future:2050-2100).

### Methods

Multi-channel singular spectrum analysis (MCSSA)^31,32^ is a multivariate version of singular spectrum analysis (SSA), a nonparametric method for the decomposition of time series into major orthogonal components. In SSA, these components are the eigenvectors of a covariance matrix constructed from a given times series against itself at different lags (for a total number of prescribed lags). The amount of variance corresponding to specific the components (including cyclical ones which come in pairs) is indicated by the corresponding eigenvalues of the eigenvectors. Components can correspond to trends, to cycles (of different frequencies, including seasonality and interannual variability) and noise. Once the major components are identified, a reconstruction of the temporal pattern of variation can be obtained from a selected set. Thus, such reconstructed time series allows selection of a range (or a specific) time scale, as well as the removal of noise and trends.

We used the SDC analysis^30, 45^ to study the local and transient patterns of variability, in particular the local correlations existent between different variables. Specifically, the two-way SDC (TW-SDC) technique computes non-parametric Spearman rank correlations between two time series in localized windows of time, using rcorr (https://www.rdocumentation.org/packages/Hmisc/versions/4.2-0/topics/rcorr). TW-SDC are computed at a variety of window sizes, S, and at different lags between the two variables. A correlation value is obtained at a given significance level (p< 0.05 or 0.01, as indicated). The SDC method is optimal for correlations between short and noisy time series, and because it is sensitive to window size, different S values, above and below the period of interest, are applied to assess for consistency in the results.

## Supplementary Figures

**Fig. S1:**
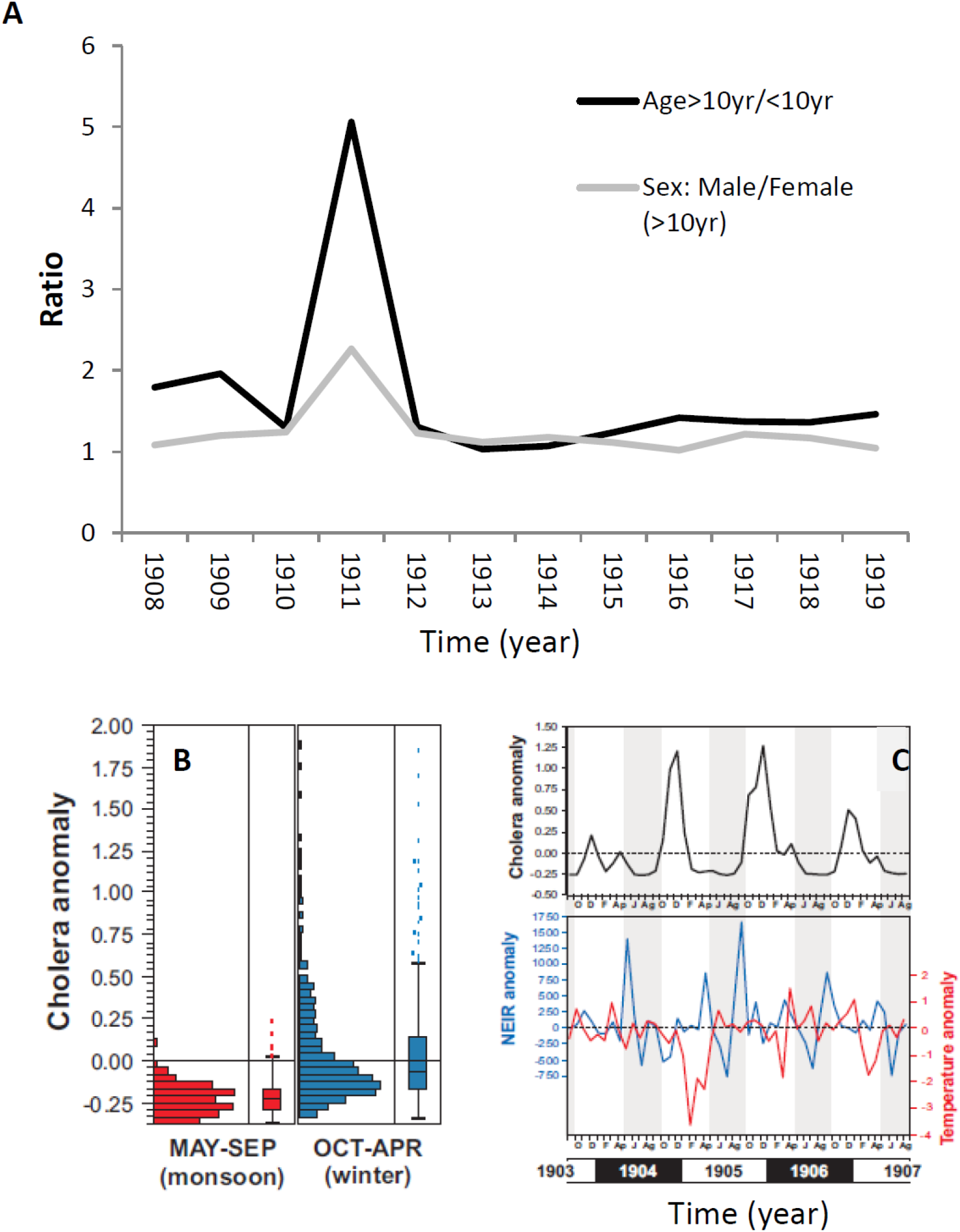
A) Ratio of Male to Female cholera deaths > 10 yr (grey line), and ratio > 10 yr of age to 10 yrs of age and below (black line). B) distribution of total occurrences of cholera mortality anomalies during the monsoons (May to September) and in winter (October to April); C) evolution of cholera mortality (anomaly), rainfall (10^−1^mm/month) and temperature (deg °C) during the anomalous 1904-07 event (grey background stripes correspond to the monsoon period).

**Fig. S2:**
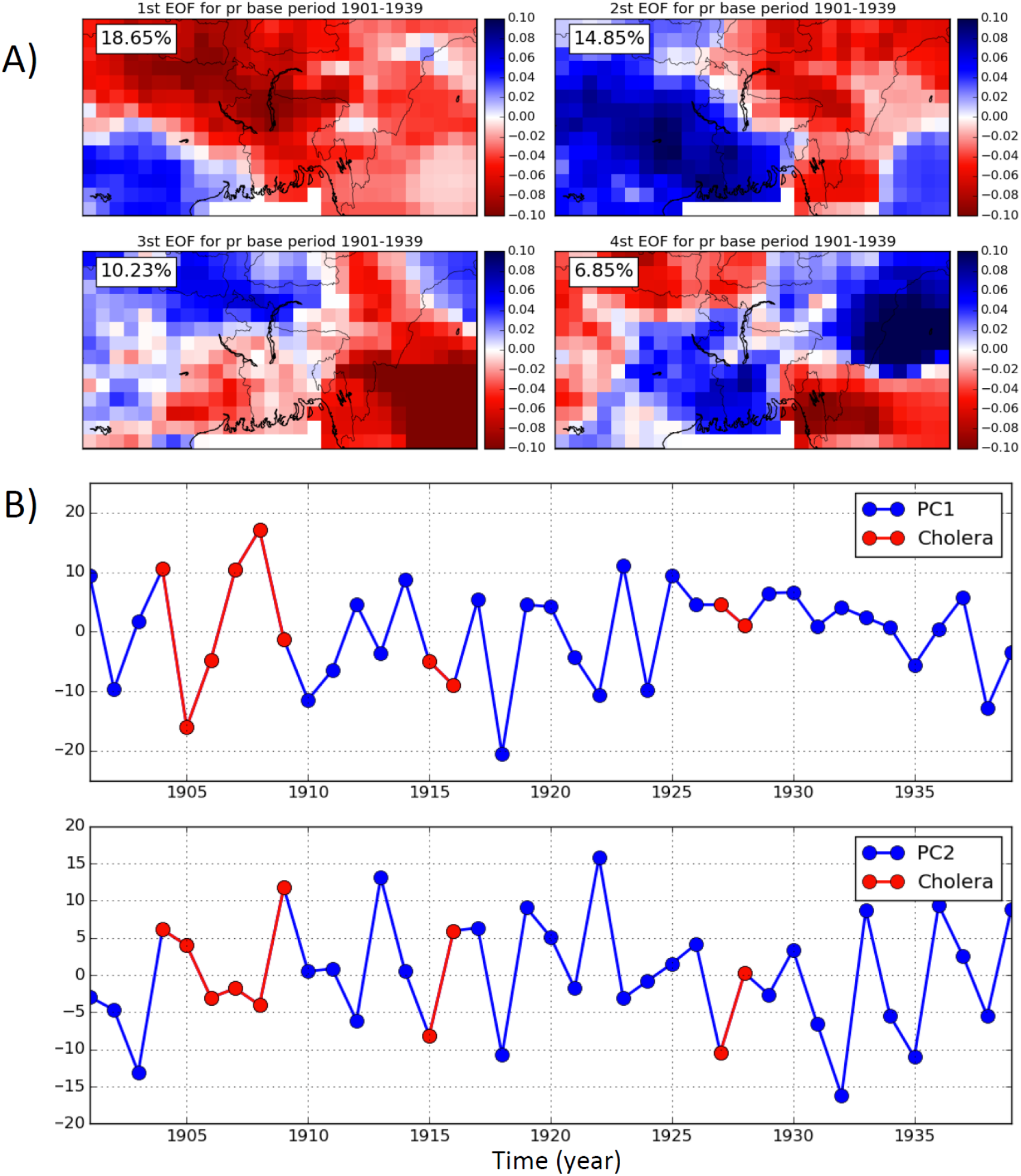
A) Principal Component Analysis (PCA) of the GPCC rainfall reanalysis^25^ over Bangladesh for the interval 1901-1940. Years before 1901 were not available. Empirical Orthogonal Functions (EOF) are significant at the p<0.01 level. B) Temporal PCs (t-PC) for the EOF components in A) with red years denoting large cholera anomalies in Bengal (*note that the sign is arbitrary in PCA*).

**Fig. S3:**
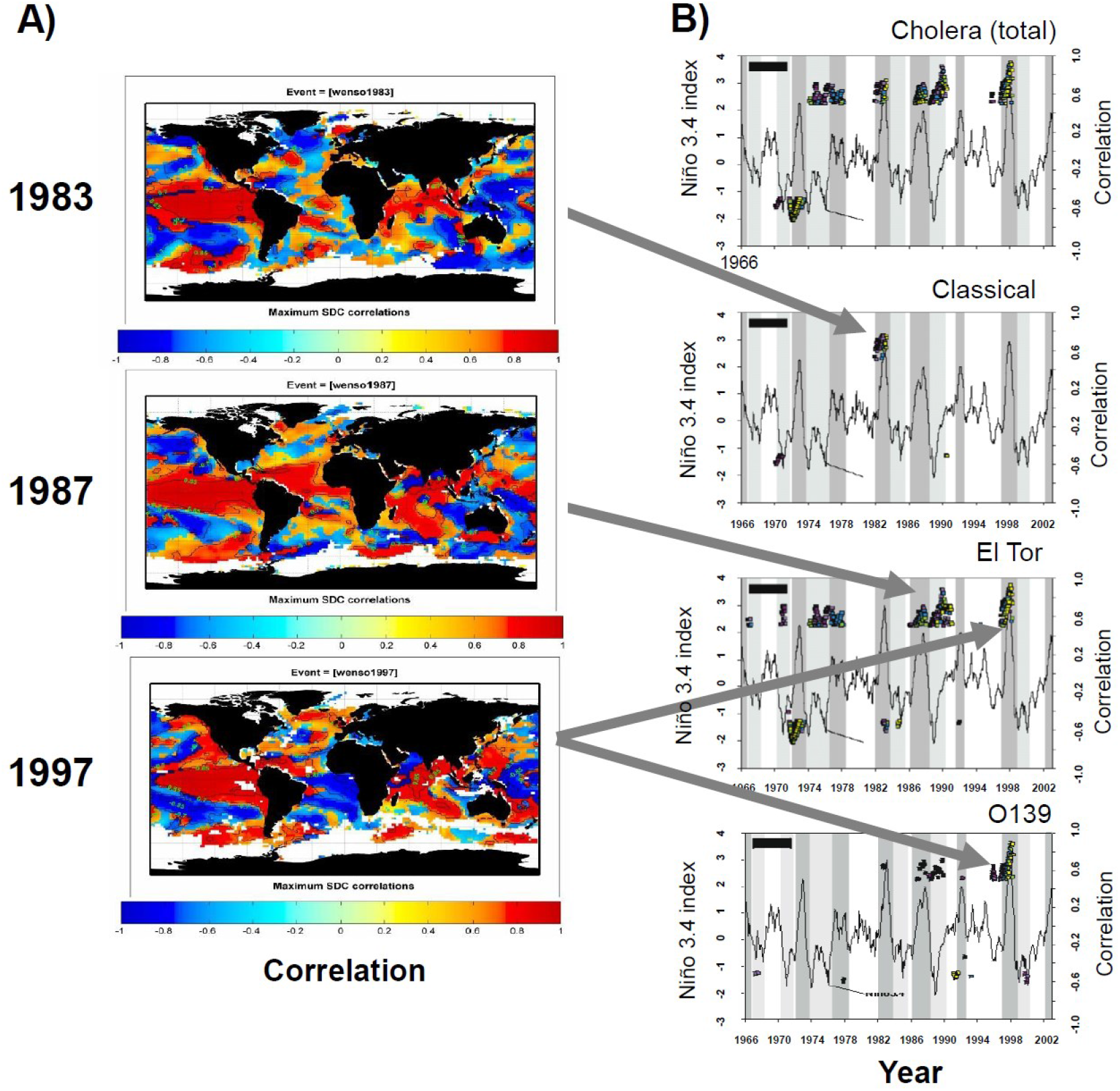
A) Scale-dependent correlation (SDC)^29^ analyses maps between sea-surface temperatures (SST) anomalies and ENSO (Niño 3.4). B) SDC analysis between total cholera cases for each of the three different strains in Bangladesh (*Classical, El Tor* and *O139*), and El Niño (*Niño3*.*4 index*), with a lead time of the climate variable of 7 to 12 months. Depicted spatial correlations correspond to the correlation maximum attained at the times when epidemics for each strain occurred in Bangladesh and the spatial replacement is known to have occurred (1982 for *Classical*, 1986-88 for *El Tor* and 1995-97 for *O139*). Lag values are indicated as coloured squares. *(panels for Classical and El Tor are adapted with permission from Pascual et al*., *Science 2000)*. Correlations were calculated locally in time for fragments of 25 months, a window size known to trace well ENSO effects^33^.

**Figure S4:**
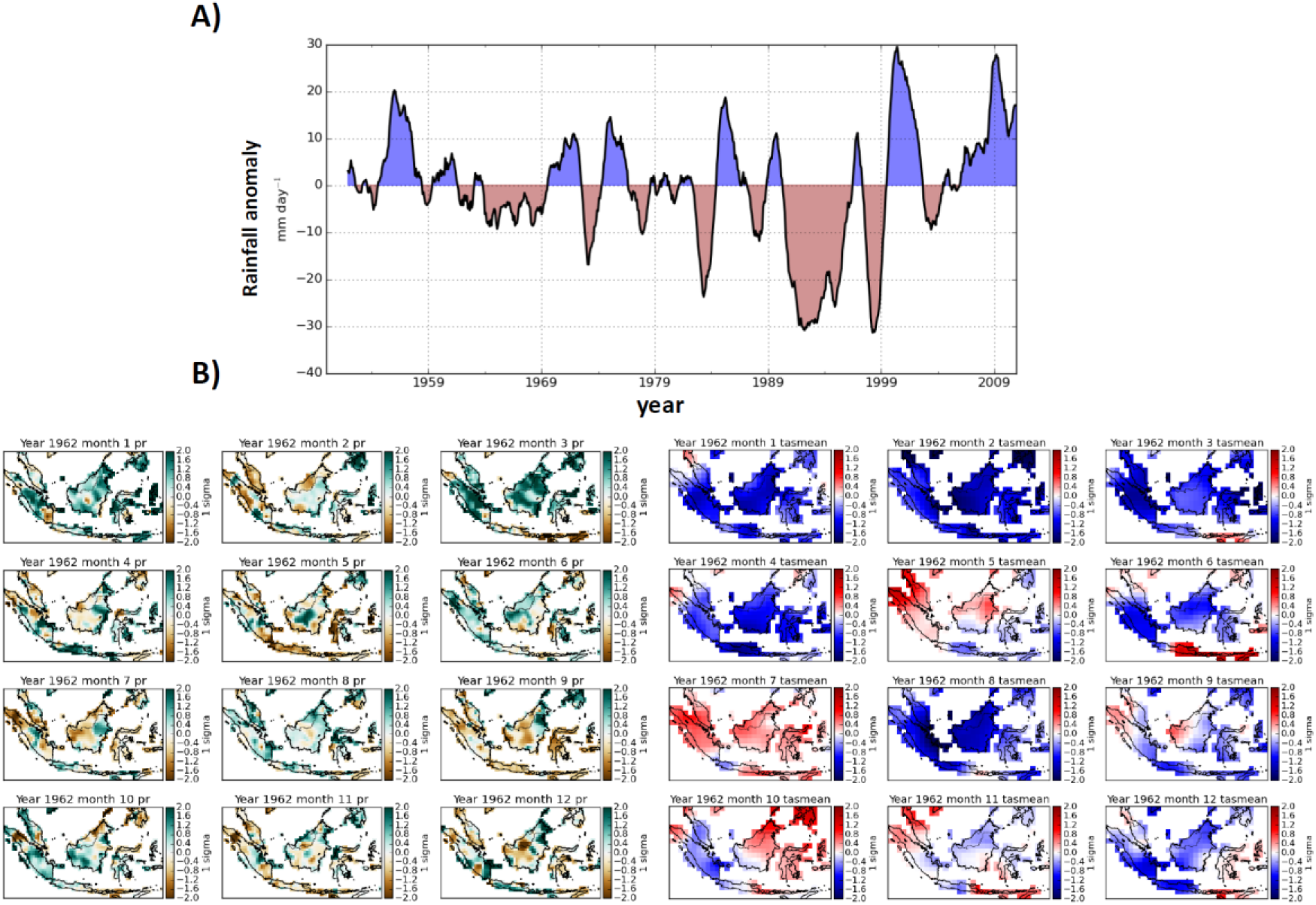
A) Time series of precipitation anomaly(*mm/day*) for the Indonesian archipelago (1950-2010). B) Monthly rainfall (*three left columns*) and mean temperature (*three right columns*) anomalies (*sigma*) in 1962, when the cholera epidemics began to propagate in and out of the country reaching Bangladesh in 1963.Number denotes month (1=January, 2=February, …, 12=December), *pr* stands for precipitation and *tasmean* for average temperature in a given month.

